# Systematic identification of disease-associated 3D neighborhoods in protein structures

**DOI:** 10.64898/2026.05.29.26354366

**Authors:** Emily Nason, Sherif Gerges, F. Kyle Satterstrom, Bram L. Gorissen, Ruqi Liao, Georgia Panagiotaropoulou, Jeremy Guez, The Autism Sequencing Consortium, Konrad J. Karczewski, Mark Daly, Hilary Finucane

## Abstract

Rare variant association studies (RVAS) have identified hundreds of genes contributing to human disease, yet gene-level signals provide limited insight into the molecular mechanisms underlying pathogenicity. Missense variants, which can be mapped onto three-dimensional protein structures, offer an opportunity to gain novel mechanistic insights. Here, we develop a scalable framework for systematically mapping case and control variants onto protein structures and identifying spatially localized regions enriched for case variants. Our framework builds on the 3D Neighborhood Test (3DNT), which we recently introduced in a single-gene analysis of *ATP2B2*, and enables the genome-wide analysis of rare coding variation beyond standard gene-level approaches. We applied 3DNT across multiple large-scale datasets, including Mendelian disease variants from ClinVar, *de novo* mutations from 37,486 autism spectrum disorder (ASD) probands, and case-control exome sequencing cohorts for epilepsy and schizophrenia. We identified significant clusters in 872 genes for Mendelian disease, in 70 genes for autism, in one gene for epilepsy, and in three genes for schizophrenia. These clusters are strongly enriched for known functional sites and provide insight into both known and previously unrecognized disease genes. Our results demonstrate that scalably integrating RVAS data with protein structure predictions localizes disease-associated variation to specific functional regions and reveals a layer of disease biology that is largely invisible to standard analyses.

## Introduction

Rare variant association studies (RVAS) have identified hundreds of genes that contribute to common diseases^1–3^. RVAS are typically driven by protein-truncating variants (PTVs), a class of variation that almost always causes loss of protein product through nonsense-mediated decay. While this property makes PTVs a powerful tool for identifying disease genes, the complete loss of function caused by PTVs limits mechanistic interpretation, which thus remains a major unsolved challenge.

Missense variants, which are roughly ten times more abundant than PTVs, offer a promising route to mechanistic interpretation because they alter specific amino acid residues and can be mapped onto three-dimensional protein structures. Some damaging missense variants broadly disrupt protein function through destabilization, whereas others perturb specific local structural neighborhoods, active sites, or protein-protein interaction interfaces. When multiple disease-associated missense variants occur within the same protein, their spatial clustering can highlight relevant molecular mechanisms and enable the detection of associations in genes for which PTVs play little or no role in disease risk^4–6^.

While previous studies have investigated the spatial clustering of missense variants in disease, systematic investigation of 3D clustering in case-control data has been surprisingly limited. For Mendelian disease, a 2018 study found that pathogenic variants cluster more than expected by chance^7^ but did not identify specific significant neighborhoods. In cancer genetics, methods like HotMAPS^6^, HotSpot3D^8^, Mutation3D^9^, and 3dhotspots^10^ have identified structural neighborhoods with an excess of somatic mutations in cancer and have shown that these neighborhoods are often located at known functional sites. However, these methods cannot be applied to case-control data, a crucial limitation in analysis of germline genetics due to potential confounding by mutation rate and regional missense constraint^11^. In neurodevelopmental diseases (NDD) and autism, previous studies have revealed one-dimensional clustering of *de novo* variants^12–14^, identified individual sites with recurrent mutations^15^, defined “Essential3D” sites in 242 NDD-associated proteins by combining evolutionary and population-based data^16^, analyzed mutational clusters in protein structures of specific protein families^17^, and compared clustering in protein structures between NDD and cancer^18^, but there has been no systematic genome-wide analysis to identify 3D neighborhoods enriched for disease mutations. Methods like POINT^19^ and deMAG^20^ use the fact that pathogenic mutations cluster in 3D protein structures for variant prioritization, but they are not designed to identify the individual clusters that are significant. PSCAN is an approach for identifying spatially-defined regions enriched for case mutations^21^, but it does not use a within-gene background, and its tested regions are defined as connected components in distance-based graphs (see **Methods**). Methods such as SCANG^22^ and earlier scan statistics^23,24^ use case-control data to associate sets of variants to disease, but define neighborhoods in 1D along the genome, rather than in 3D based on protein structures. And while predictors of pathogenicity and deleteriousness such as AlphaMissense^25^ and PopEVE^26^ have impressive performance, they do not provide insight into 3D clustering.

In recent work^27^, we introduced the 3D Neighborhood Test (3DNT) in a single-gene analysis, establishing a proof of concept that spatial clustering using RVAS data alone can be used to identify disease-relevant functional regions within protein structures. 3DNT identifies spherical neighborhoods with significant enrichment of case mutations using only case/control labels. Applied to *ATP2B2*, 3DNT recovered neighborhoods overlapping the calcium binding site and the ATP:Mg²⁺ cofactor binding site, and functional experiments confirmed that variants from both neighborhoods disrupted calcium transport. This success motivated the development of a general and computationally efficient framework that could extend this single-gene proof of concept to a genome-wide analysis across multiple diseases.

#### Box 1. Outline of the 3D neighborhood test. R is a parameter set by the user.

1. Map case and control variants to protein structure

2. For the neighborhood of radius R centered around each residue: Compare case:control ratio in neighborhood to case:control ratio in rest of protein with Fisher’s exact test

3. Permute case/control labels and compute empirical FDRs

Here, we develop a scalable framework based on 3DNT and apply it across multiple large-scale datasets: Mendelian disease variants from ClinVar^28^, *de novo* variants from probands with autism spectrum disorder (ASD)^29^, and case-control exome sequencing datasets for schizophrenia^3^ and epilepsy^30^. We find widespread significant clusters of disease variants, which provide insights into potential molecular mechanisms, highlight specific high-confidence missense variants for these challenging diseases, and identify disease genes with non-haploinsufficient mechanisms, demonstrating that, at modern sample sizes, systematic integration of case-control genetic data with protein structure prediction enables insights that are invisible to standard gene-based analyses.

### Overview of method

Our method tests spheres of fixed or variable radius in the 3D predicted protein structure for an excess of case mutations vs control mutations. To do this, we first map input mutations to the protein structures, and we construct a sphere around each residue in the protein. Our default radius is fixed at 15 angstroms, which is the smallest radius at which we have acceptable power, but analysis with different or multiple radii is also enabled; for a discussion of radius selection, see **Methods**. For each sphere, we then use Fisher’s exact test to compare the ratio of case mutations to control mutations in the sphere vs. in the remainder of the protein. We use permutation of the case-control labels to compute an empirical false discovery rate (FDR) and family-wise error rate (FWER) over all residues in all proteins included in the analysis (**Methods**, **Box 1**). We note that such testing is competitive within the gene and so provides significance estimation of the structure-based clustering alone, independent of any difference in the relative rates or counts of the case and control variants overall. Because of the computational expense of counting mutations and running Fisher’s exact test for every residue in every protein in an analysis, we introduce several computational innovations that result in a many-fold speed increase over a naive implementation, without compromising accuracy (**Methods**). The run time of our method depends on the number of mutations and the size of the protein, but, after a few seconds to load the reference files, runs in less than a second per protein on a standard laptop for most proteins analyzed in this paper. Our method is freely available at github.com/FinucaneLab/structure-informed-rvas.

In this work, we describe results using parameter settings which drop common variation (allele count greater than five in the data set under study, or allele frequency greater than 0.001 in any population in gnomAD) to avoid the effects of linkage disequilibrium and to take advantage of the larger average effect sizes of ultra-rare variants.

### 3D neighborhoods enriched for pathogenic variants in ClinVar

We first applied 3DNT to interpret Mendelian disease variants from ClinVar. Previous work has shown that Pathogenic / Likely pathogenic (P/LP) variants in ClinVar cluster in 3D space, on average, more than variants observed in gnomAD^7^, and previous analyses of individual proteins have found cases where P/LP variants cluster near known functional sites^31^. However, to our knowledge, no previous analysis has systematically scanned all ClinVar data to identify the significant clusters of P/LP variants in protein structures.

To identify clusters of P/LP variants in ClinVar, we applied 3DNT, assigning the “case” label with allele count of one to Pathogenic / Likely pathogenic (P/LP) variants and the “control” label with allele count of one to Benign / Likely benign (B/LB) variants. Variants in gnomAD that were common in at least one population were added as additional controls, and the default behavior of dropping common variants was overridden. Of 1,121 proteins with at least 5 case variants and 5 control variants, 872 had at least one neighborhood that reached FDR < 0.05 in this analysis, and 176 had at least one neighborhood that reached FWER < 0.05, suggesting the widespread utility of this approach (**Fig. 1a**). We found that significantly more genes associated with autosomal dominant disorders harbored significant neighborhoods at FWER < 0.05 than genes associated with autosomal recessive disorders (137/604 vs 45/655; P=9e-16), consistent with previous literature on clustering of pathogenic variants in these two classes of genes^32^.

**Figure 1.**
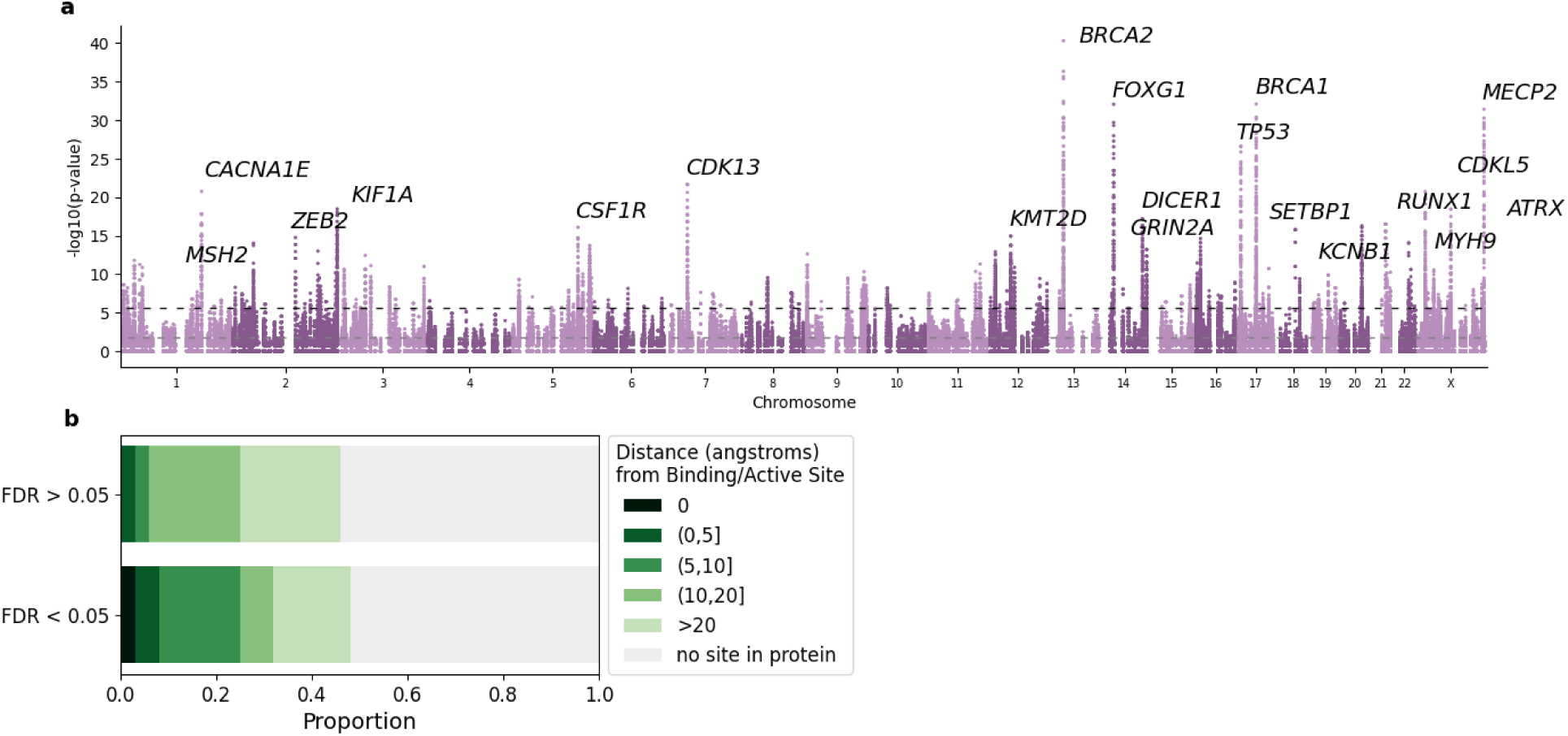
Results of applying 3DNT to data from ClinVar. **(a)** Significance of each neighborhood in ClinVar. Grey dashed line indicates FDR = 0.05; black dashed line indicates FWER = 0.05. **(b)** Comparison of the distances of significant vs non-significant neighborhood centers to UniProt-annotated binding/active sites, with one neighborhood per gene (**Methods**).

We characterized these results using UniProt-annotated binding and active sites, AF2Bind-predicted binding sites^33^, and relative solvent accessible surface area. To compute significance of enrichment given the correlation among nearby neighborhoods, we compared the centers of the most significant neighborhoods in proteins with FDR < 0.05 to random neighborhoods in other proteins, restricting to one neighborhood per protein. The centers of the most significant neighborhoods were closer to UniProt-annotated binding/active sites, closer to AF2Bind-predicting binding sites, and more likely to be buried than random neighborhoods (P =1.6e-6, 9.83-9, and 4.4e-12, respectively; see **Methods** and **Fig. 1b**). While ClinVar P/LP variants are more likely than B/LB variants to be found in the hydrophobic core of proteins (OR=6.7, P=<1e-15), we found that the proportion of P/LP variants in significant neighborhoods that were in the hydrophobic core was not significantly different from the proportion of P/LP variants in randomly selected neighborhoods that were in the hydrophobic core (P=0.39).

### 3D neighborhoods enriched for de novo variants in autism probands

To our knowledge, there has been no previous genome-wide scan to identify 3D neighborhoods with an excess of *de novo* variants in autism. We applied 3DNT to an exome sequencing data set of autism probands and their parents (N=37,486 probands), analyzing the 800 genes with more than five *de novo* missense variants in autism probands after mapping to predicted structures. Of these 800 genes, 144 were associated to ASD at FDR < 0.001 in the flagship analysis^29^. Within these genes, we assigned the “case” label to the *de novo* missense variants in autism probands with allele count equal to the number of carriers, and the “control” label to singleton untransmitted alleles found in the parents of those probands in the same gene.

Overall, 70 / 800 genes had at least one significant neighborhood at FDR < 0.05, of which 36 genes had been identified as autism-associated (at FDR < 0.001) in the flagship analysis. Nine genes had at least one significant neighborhood at FWER < 0.05, of which seven had been identified as autism-associated in the flagship analysis (**Fig 2a-c**). Among FDR-significant neighborhoods, 17% were centered within 10Å of UniProt-annotated active or binding sites, representing a 9-fold enrichment over non-significant neighborhoods (**Fig 2b**). The most significant neighborhood, in *CDK13*, was directly at the catalytic pocket, recapitulating previous observations^34^. The most significant neighborhood in the next protein, encoded by *DEAF1*, was in the SAND domain, which has previously been shown to be functional and harbor disease mutations^35^. The third protein was encoded by *KCNQ3*, in which the most significant neighborhood harbored nine case mutations at six sites; eight of the nine were mutations from arginine in the voltage sensing domain. Previous work has shown that positively charged residues in the voltage sensing domain are crucial for the function of potassium channels^36^. Moreover, the R230C mutation, which occurs three times in probands in this data set, has been previously found in individuals with autism and shown to have a gain-of-function effect^37,38^.

**Figure 2.**
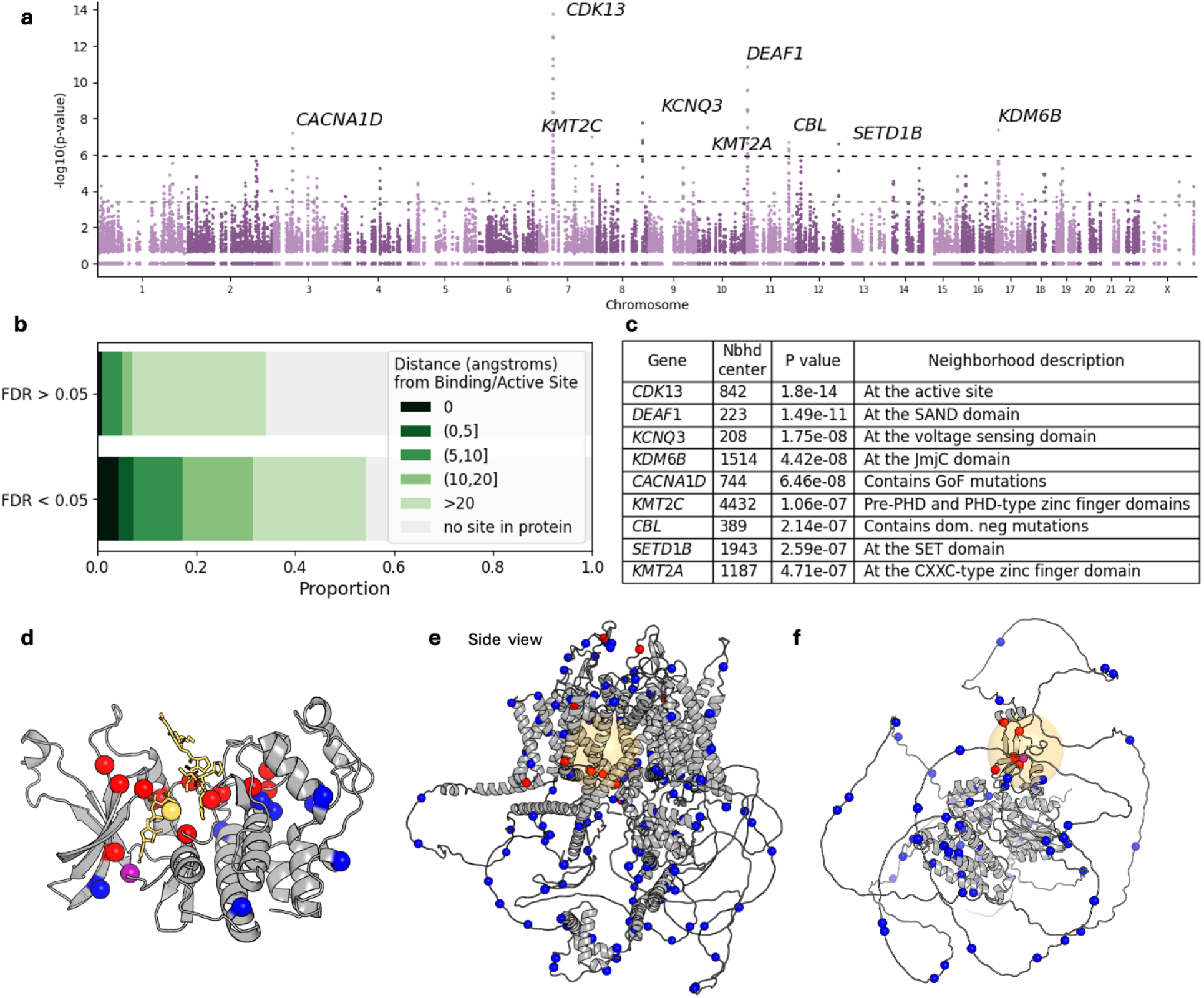
Results of applying 3DNT to *de novo* variants from 37,486 autism probands, with data from the Autism Sequencing Consortium, restricting to genes with at least five *de novo* missense variants. **(a)** Significance of each neighborhood tested. Grey dashed line indicates FDR = 0.05; black indicates FWER = 0.05. **(b)** Comparison of the distances of significant vs non-significant neighborhoods to UniProt-annotated binding/active sites, with one neighborhood per protein. **(c)** Table of the top neighborhoods in the nine genes for which the top neighborhood reaches FWER < 0.05. **(d)** Predicted structure of the kinase domain of *CDK13*, with ATP, Mg^2+^, and peptide substrate added via alignment with PDB 1QMZ. Variants in the ASC dataset are shown as spheres. Red, case-only; blue, control-only; purple, residue harbors both case and control variants. **(e)** AlphaFold-predicted structure of CaV1.3, encoded by *CACNA1D*. Residues blocking the view of the top neighborhood (14% of all residues) have been hidden. Orange ball indicates the most significant neighborhood. Case and control variants are displayed as in (d). **(f)** AlphaFold-predicted structure of *CBL*. Case and control variants are displayed as in (d).

The nine genes with neighborhoods at FWER < 0.05 also included two genes not identified in the flagship analysis: *CACNA1D* and *CBL*. In *CACNA1D*, a calcium channel (**Fig 2d**), five of twelve *de novo* missense mutations in probands lie in a 15 angstrom ball that covers 3% of the protein. The neighborhood is located near the pore, at the boundary of the transmembrane domain and the cytoplasmic domain. Two of the five mutations in this neighborhood, A749G and G407R, have been previously reported in probands with autism and shown to be gain-of-function mutations^39^. In the E3 ubiquitin ligase CBL, the most significant neighborhood covers 6% of the protein and contains 6 / 6 *de novo* variants in probands and 0 / 66 control variants (**Fig 2e**). This neighborhood overlaps the linker-RING finger domain, and it is also highly significant in the analysis of data from ClinVar (13/15 P/LP variants, 0/19 B/LB variants, FWER <1e-3 with 1,000 simulations). *CBL* is a known disease gene with phenotypes including developmental delay, intellectual disability, and increased risk of juvenile myeloid leukemia. The Q367P mutation, identified here as a *de novo* variant in an autism proband, was previously shown in a study of mutations in myeloid neoplasms to abolish E3 ubiquitin ligase activity in a dominant-negative fashion, while also producing gain-of-function effects through enhanced cytokine sensitivity^40^.

The four remaining genes with significant neighborhoods at FWER < 0.05 were the lysine demethylase encoded by *KDM6B* and the histone lysine methyltransferases encoded by *SETD1B*, *KMT2C,* and *KMT2A*. KDM6B removes the repressive mark H3K27me3, and the top neighborhood overlaps the JmjC domain, which harbors the catalytic center for this protein. SETD1B, KMT2C, and KMT2A deposit activating H3K4 methylation; the top neighborhood in SETD1B overlaps the catalytic SET domain, while the top neighborhoods in KMT2C and KMT2A overlap zinc finger domains which are responsible for localization and regulation of catalytic activity.

### Increasing power for case-control cohorts

In a case-control study, unlike the ClinVar and autism trio data sets analyzed above, all ultra-rare missense variants carried by cases are treated as case variants. Because most such variants are inherited—and *de novo* variants cannot be distinguished without trio data—the resulting set is dominated by inherited variation, which contains a substantially higher proportion of non-causal variants. As a result, the case variant set is markedly less enriched for causal mutations than in *de novo* studies, placing the analysis in a more challenging power regime. However, even in this regime, we hypothesized that it would be possible to identify 3D neighborhoods with an excess of case mutations if the truly causal mutations exhibit clustering.

To increase power in this challenging setting, we imposed two filters. First, we analyzed only genes with some *a priori* genetics-based evidence of association to the phenotype: either a missense-only burden p-value more significant than 0.001, or an overall association in the flagship analysis passing FDR < 0.05. Although restricting the analysis in this way limits the potential novelty of the results, any significant neighborhood identified would strengthen evidence for the gene beyond that provided by P < 0.001 and/or FDR < 0.05, while also providing mechanistic and variant-level insights.

Second, we sought to pre-identify a subset of neighborhoods more likely to be functional while avoiding the bias introduced by human-curated annotations. To do this, we used AlphaMissense^25^ (AM) pathogenicity scores—which are derived from protein sequence and structural context, with no input from case-control disease data—to identify structurally and evolutionarily constrained neighborhoods across the proteome. Specifically, we treated positions with high AM scores as “case” variants and positions with low AM scores as “control” variants, then applied 3DNT genome-wide to identify neighborhoods with a significant excess of high-AM positions (**Methods**). Due to the strong spatial patterning in AM scores, over half of all neighborhoods proteome-wide had FDR < 0.05 in this analysis. The set of neighborhoods with FDR = 0 (i.e., a raw p-value more significant than that generated in any of the 1,000 null simulations) constituted 15% of all neighborhoods genome-wide, with 93% of proteins containing at least one neighborhood in this set. To increase power in our analysis of the case-control datasets of interest (schizophrenia and epilepsy), we therefore analyzed only neighborhoods which had an FDR of 0 in this AM analysis, rather than all neighborhoods in all genes. Because AM scores share no information with the disease case-control labels used in 3DNT, this filter introduces no circularity: neighborhoods are still tested exactly as in the original method, with a Fisher’s exact test comparing the case/control ratio in the neighborhood to the ratio in the remainder of the protein. The only limitation introduced is that neighborhoods outside the “AM filter” set are not considered. While this restricts the set of discoverable neighborhoods, it does so in a way that increases power, retains a large fraction of the proteome, and avoids reliance on human-curated functional annotations.

### 3D neighborhood enriched for case mutations in a case-control cohort for epilepsy

We analyzed a public case-control exome sequencing data set for epilepsy from the Epi25 consortium^30^ (N_case_=20,979; N_control_=33,444). In this data set, there are eight genes with missense-based burden p-value < 0.001 and three genes with a PTV-based burden FDR < 0.05. We analyzed these 11 genes using the AM filter described above. In this analysis, one neighborhood in KCNA1 reached FWER < 0.05 (**Fig 3a**). This neighborhood included 8 case variants and zero control variants, with two of the case variants overlapping the highly conserved selectivity filter motif TVGYG (**Fig 3b-c**). Both of the case variants that overlapped the selectivity filter were mutations from the small, nonpolar amino acid glycine to the large, positively charged amino acid arginine; both would thus be expected to cause a large disruption to this highly conserved and functionally critical unit of the protein.

**Figure 3.**
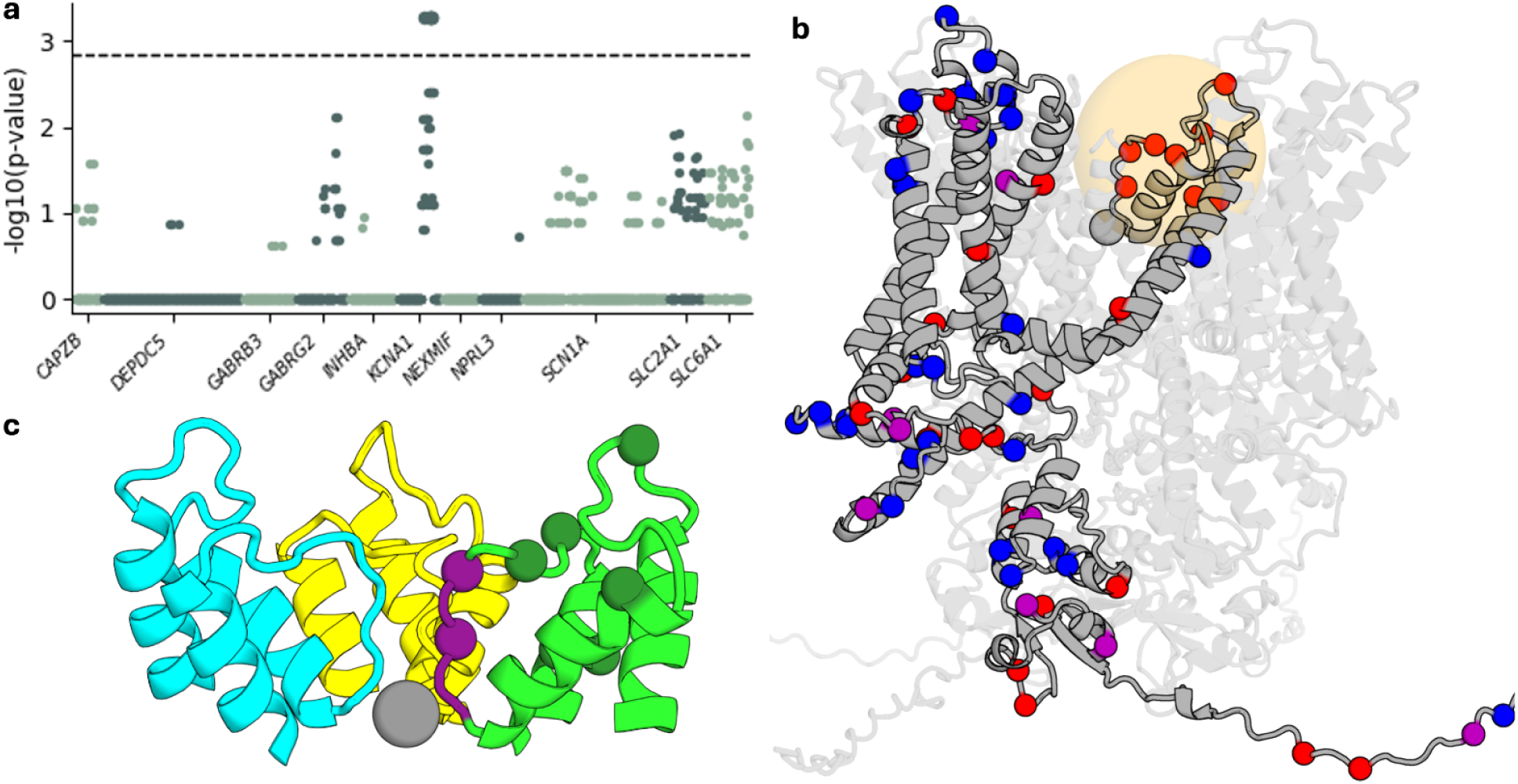
Analysis of data from the Epi25 study. (a) Significance for all neighborhoods in the analysis. Dotted line indicates the FWER = 0.05 and FDR = 0.05 cutoffs, which are shared in this analysis. P-values greater than 0.05 are not computed precisely for computational reasons, and are reported as one. (b) AF3-predicted structure of the KCNA1/KCNA2 tetramer, with K^+^ ion. Variants in the Epi25 dataset are shown as spheres on one of the KCNA1 subunits. Red, case-only; blue, control-only; purple, residue harbors both case and control variants. Orange ball, significant neighborhood. (c) AF3 prediction of the structure of the selectivity filter region of the KCNA1/KCNA2 tetramer, with K^+^. Green and cyan, the two KCNA1 subunits. Yellow, one copy of KCNA2 (second copy omitted for visualization purposes). Purple, selectivity filter of one KCNA1 subunit. Grey, K^+^ ion. Spheres, case variants in the top neighborhood. There are no control variants in the neighborhood.

To evaluate the potential impact of one of these case mutations, G374R, we used FoldX^41^, a physics-based predictor of mutational effect, applied to an AlphaFold3-predicted structure of the KCNA1/KCNA2 heterotetramer in complex with K⁺. The mutation was predicted to markedly reduce binding affinity between KCNA1 and KCNA2 subunits (predicted ΔΔG > 25 kcal/mol), consistent with substantial destabilization of heterotetramer formation. This result is consistent with experimental data from the homologous protein KcsA showing that destabilization of tetrameric assembly is a common consequence of mutations in the selectivity filter^42^.

### 3D neighborhoods enriched for case mutations in a case-control cohort for schizophrenia

Next, we analyzed public data from a case-control exome-sequencing study of schizophrenia from the SCHEMA consortium^3^ (N_case_ = 24,248; N_control_ = 97,322). We applied 3DNT to the set of 32 genes with FDR < 5% in the flagship analysis and the five genes with missense-based burden P<0.001, again using the AM filter. We found that three genes, *GRIA3*, *SETD1A,* and *ATP2B2*, had neighborhoods with FDR < 0.05, with *GRIA3* and *SETD1A* neighborhoods passing the more stringent FWER threshold (**Fig 4a**). The significant neighborhood in GRIA3, a glutamate receptor, was adjacent to the L-glutamate binding site (**Fig 4c-e**). The significant neighborhood in *ATP2B2* was the neighborhood that inspired the development of this method and is discussed in detail in ref [27], where its significance is further augmented by the combination of data from both schizophrenia and autism cohorts.

**Figure 4.**
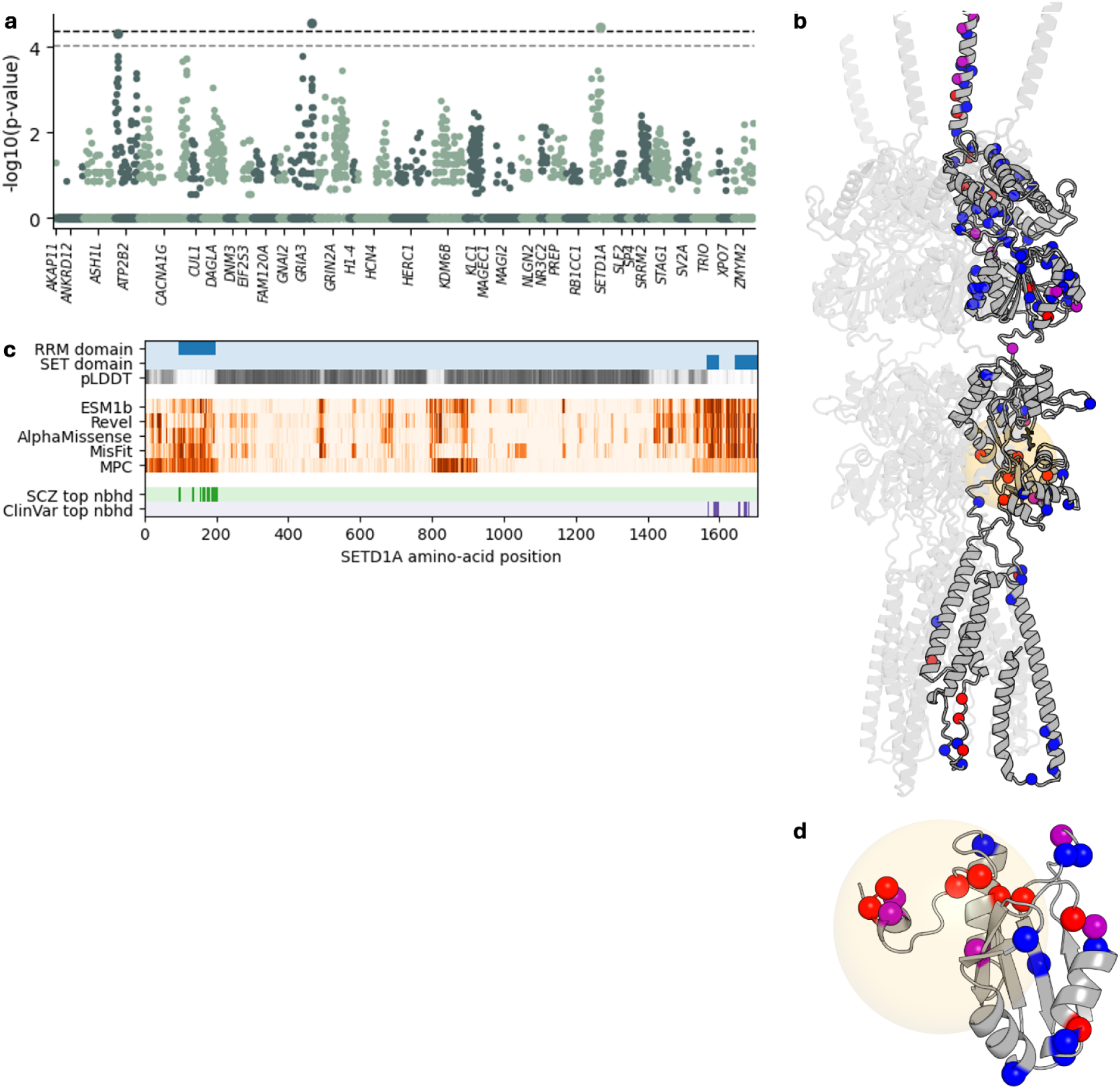
Analysis of data from the SCHEMA study. (a) Significance for all neighborhoods in the analysis. Grey dotted line, FDR = 0.05; black dotted line, FWER = 0.05. P-values greater than 0.05 are not computed precisely for computational reasons, and are reported as one. (b) Predicted structure of the *GRIA3* homotetramer (with signal peptide included). L-glutamate added by alignment to PDB CDLN and shown in black. Variants in the SCHEMA dataset are shown as spheres on one of the subunits. Red, case-only; blue, control-only; purple, residue harbors both case and control variants. Orange ball, significant neighborhood. (c) Top, locations of the RRM and SET domains of SETD1A, as defined in The Encyclopedia of Domains (TED) 49 . Middle, percentiles of five missense variant impact predictors, on average over mutations at a position. Bottom, location of the most significant neighborhoods in SCHEMA and ClinVar. (d) Predicted structure of the RNA recognition motif (RRM) domain. Colors as in Fig (b).

The most significant neighborhood in *SETD1A* was in the RNA recognition motif domain (**Fig 3f**). SETD1A is a histone methyltransferase that catalyzes H3K4 methylation. It contains a well-characterized SET domain at the C-terminus that harbors the catalytic site, and an RNA recognition motif (RRM) domain at the N-terminus, with a largely disordered region (low pLDDT) between them. Five missense variant effect predictors^11,25,43–45^ predict that both the RRM and SET domains are intolerant to mutation, with the RRM domain showing particularly strong constraint signal from MPC^11^, which captures regional missense depletion in human populations. The significant 3DNT neighborhood in SETD1A overlaps the RRM domain. Notably, the yeast homolog Set1 also contains an RRM domain that is required for H3K4 trimethylation but not for dimethylation^46,47^. While the functional role of the RRM domain in human SETD1A remains less well characterized, SETD1A has been shown to bind RNA^48^. *SETD1A* also has a neighborhood that reaches FDR < 0.05 in ClinVar; this neighborhood overlaps the SET domain. The most significant neighborhood in ClinVar is non-significant in SCHEMA, and the most significant neighborhood in SCHEMA is non-significant in ClinVar.

### Comparison to other methods

Although no other method we are aware of tests 3D neighborhoods for an excess of case mutations compared to a within-gene background proportion of case mutations in a case-control study, there are several methods with related goals. We thus sought to compare their performance on the four data sets analyzed here.

PSCAN^21^ is the most closely analogous method to 3DNT: it takes as input case and control mutations, maps them to 3D protein structures, and reports significant neighborhoods in 3D space. Unlike 3DNT, which performs a competitive test against the within-gene background proportion of case mutations, however, PSCAN tests neighborhoods for disease association without comparison to the within-gene background. PSCAN also tests a set of spatially defined neighborhoods that is very different from the set of neighborhoods tested by 3DNT (**Methods**). We repurposed PSCAN as a competitive within-gene test, so that it would not report significant neighborhoods in a gene with an excess of case missense variants but no spatial clustering, and we applied it to the four data sets analyzed here (**Methods**). PSCAN identified far fewer significant genes than 3DNT across all four data sets: 25 vs. 872 in ClinVar, 1 vs. 70 in ASD, 1 vs. 3 in SCHEMA, and 0 vs. 1 in Epi25 (**Figure 5a**). The results also had weaker enrichment for annotated binding and active sites than 3DNT (**Figure 5b**), and neighborhoods were larger than 3DNT-identified neighborhoods (**Figure 5c**).

**Figure 5.**
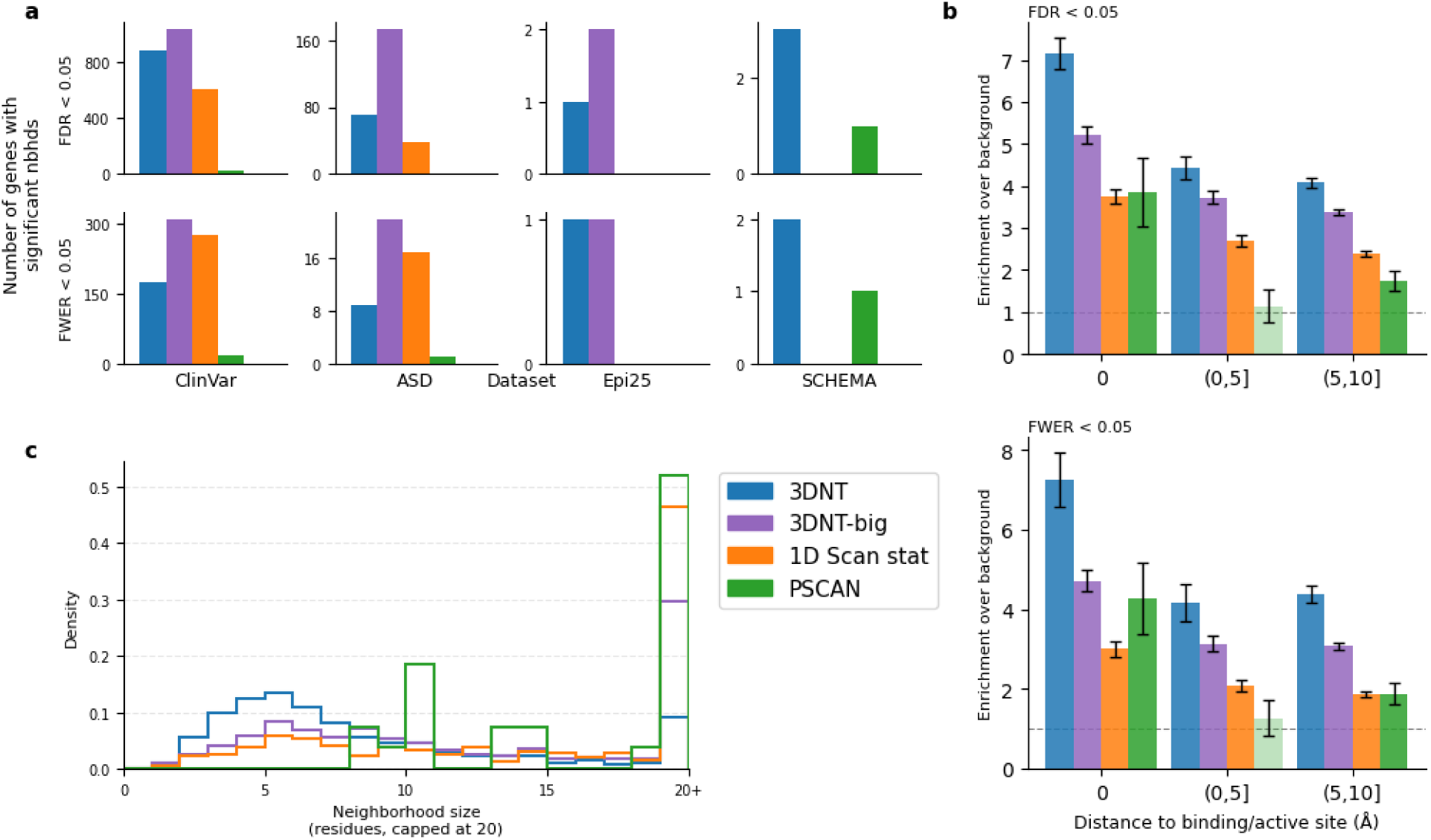
Comparison to other methods. (a) Number of genes identified in the four data sets by each of the four methods. Note the different y axes for different data sets. (b) Enrichments of residues in significant neighborhoods for being close to annotated active and binding sites from UniProt. For each gene with a significant neighborhood at FDR (top) or FWER (bottom) < 0.05, all residues in the most significant neighborhood are included. (c) For each gene with a significant neighborhood at FDR < 0.05, the total number of case and control variants in the most significant neighborhood.

POINT^19^ is a method that conducts, for each index variant, a set test in which variants are weighted according to their distance to the index variant using a Gaussian kernel. As with PSCAN, POINT compares each set to the null of no association to disease, rather than a per-gene background, but can be adapted to find spatial signals by including only data from the gene. Although POINT is described as a method for prioritizing variants, it can be interpreted as an association test for neighborhoods, analogous to 3DNT. We adapted POINT to apply to our data, but it did not find significant results, likely because it was not designed for application to data in which all allele counts are between 1 and 5 (**Methods**).

There are several methods designed to identify clusters of disease variants in 1D along the genome, without mapping to protein structures^22–24^. To analyze the summary statistics considered here, we implemented the 1D scan statistic described in ref [^23^] and applied it to our four data sets. Because of the large multiple testing burden and the low resolution of permutation-derived p-values, we adapted the empirical FWER/FDR from 3DNT for use with the 1D scan statistic. In ClinVar and ASD, the 1D scan statistic found more results at FWER < 0.05 and fewer results at FDR < 0.05 than 3DNT. It found no significant neighborhoods in Epi25 and SCHEMA (**Figure 5a**). Similar to PSCAN, the neighborhoods identified by the 1D scan statistic were larger than those identified by 3DNT and had weaker enrichment for annotated active and binding sites (**Figures 5b,c**). We note, though, that 1D neighborhoods may be enriched for other types of functional elements, including elements such as degrons that are not as well-annotated in UniProt as active sites.

To emulate the performance of the 1D scan statistic in finding a larger number of larger neighborhoods with less biological interpretability, we introduced an alternate version of 3DNT in which the test is performed at multiple neighborhood radii (10, 15, 20, 30, and 40 angstroms) and the p-values from the different radii are combined through the Harmonic Mean P-value method^50^ at each residue. We call this method 3DNT-big. Compared to 3DNT, 3DNT-big identifies more genes with significant neighborhoods in the ClinVar and ASD data sets, but the neighborhoods are larger and less enriched for active and binding sites. Compared to the 1D scan statistic, though, 3DNT-big identified more significant results with higher functional enrichment, with smaller neighborhoods on average (**Figure 5**). Our focus in this manuscript is on biological interpretability, and 15 angstroms is the smallest radius at which we expect to be powered to identify significant signals (**Methods**). We thus maintain a fixed radius of 15 angstroms as our default.

In addition to these methods, there are a number of methods designed to identify clusters of somatic mutations in cancer data^6,8–10^. However, these methods operate only on disease mutations, without control mutations for comparison. In germline genetics, because of mutational hotspots and regional missense constraint^11^, the distribution of case-only mutations can be highly non-uniform without any disease enrichment. Thus, we focus on methods designed for analysis of case-control data.

## Discussion

In this study, we develop a scalable and statistically rigorous framework for systematically mapping disease-associated variation onto three-dimensional protein structures and identifying compact structural regions enriched for pathogenic missense variants across the proteome. By moving beyond gene-level analyses to a within-gene competitive analysis, this approach allows genetic associations to be resolved to specific functional regions within proteins, providing a direct link between rare variation and molecular mechanism.

Applying this framework across multiple large-scale datasets, we identify numerous significant 3D neighborhoods across diverse classes of disease genes, including those driven by haploinsufficiency, gain-of-function, and dominant negative mechanisms. Even in less well-powered analyses of inherited variation, we detect functionally informative clusters, such as in *KCNA1* for epilepsy and in *GRIA3* and *SETD1A* for schizophrenia, that highlight specific variants and regions for experimental follow-up. To our knowledge, this work represents the first large-scale, systematic identification of compact three-dimensional clusters of germline disease variation across the human proteome.

Our method comes with several limitations. First, it is designed for ultra-rare variants and does not account for linkage disequilibrium. Extending this approach to more common variation will require incorporating LD, likely through in-sample LD computation or individual-level data. In addition, although single multi-nucleotide mutational events called as multiple missense variants are rare, they may inflate the test statistic when they occur. Second, the use of spherical neighborhoods enables detection of compact functional regions but may miss more complex spatial patterns. For example, protein-protein interaction interfaces and other extended structural features may not be well captured by this approach, motivating future methodological development tailored to these elements. Additionally, linear sequence features such as degrons may be more effectively detected by a 1D scan statistic than by 3DNT; the complementarity of 1D and 3D analyses remains an area for future research. Third, power remains limited in case-control exome sequencing studies such as those for schizophrenia and epilepsy analyzed here. Incorporating additional information such as biochemical properties of amino acids may improve sensitivity in these settings. Finally, while the neighborhoods we identify are enriched for disease association, follow-up studies are required to unambiguously implicate individual variants in disease.

As sequencing sample sizes grow and protein structure prediction continues to improve, integrating genetic data with three-dimensional protein structure is likely to become an increasingly powerful approach for interpreting rare variation. More broadly, our results suggest that systematic analysis of protein structure can complement gene-level association studies by localizing disease-associated variation within proteins, providing a systematic framework for connecting genetic association to molecular mechanism.

## Data Availability

All data produced in the present study are available upon reasonable request to the authors

## Acknowledgements

We thank Yakir Reshef, Siwei Chen, Jordan Safer, and Sumaiya Iqbal for helpful conversations. This work was supported by the Stanley Center for Psychiatric Research and the Novo Nordisk Foundation (NNF21SA0072102).

## Software availability

Our software is freely available at https://github.com/FinucaneLab/structure-informed-rvas/.

## Methods

### Use of large language models (LLMs)

We found LLM-based tools to be very helpful during this project. We used Claude Code extensively, both during development of the core software and for the analyses in the paper. While we found Claude Code to have high—and increasing—usefulness, it required close supervision and was unhelpful for the most complex parts of the software development. During manuscript preparation, we found LLM-based tools to be helpful for figure-making code, for feedback on writing, and for help clarifying and revising individual paragraphs.

### Mapping genetic coordinates of missense variants to protein structures

We began with a table of genome-wide annotations from the Variant Effect Predictor^51^ (VEP) tool, generated by the gnomAD project and publicly available at gs://gcp-public-data--gnomad/resources/context/grch38_context_vep_annotated.v115.ht. This table is based on Ensembl version 115. We included all possible missense variants on autosomes or the X chromosome, excluding those that involved non-standard amino acids.

Using tables created from BioMart in R for Ensembl 115, we mapped Ensembl transcripts to UniProt isoforms. When available, we used the isoform identified as canonical by UniProt for reviewed UniProt IDs. If none were available, we used non-canonical isoforms of reviewed proteins, and if that was not available, then unreviewed UniProt IDs. For Ensembl transcripts whose sequence matched the sequence of the UniProt isoform except for an initial ‘X’, we dropped the ‘X’ and adjusted the indexing to match the UniProt sequence. For UniProt isoforms mapped to by multiple Ensembl transcripts, we kept the transcript that had the highest sequence similarity, prioritizing transcripts labelled as “protein_coding” in Ensembl when there was a tie for highest sequence similarity. There were 95 genes that did not appear in the BioMart table and were dropped.

Following these steps, there were 34 proteins where the sequence of the Ensembl transcript did not match the sequence of the UniProt isoform. Of these, five IDs had failures that were due to a mapping error in the BioMart-generated Ensembl-UniProt table and were manually fixed. Four IDs no longer existed in UniProt and the Ensembl transcripts did not map to any other UniProt isoforms, and these were dropped from the mapping table. For the remaining 25 IDs, we used the Ensembl transcript sequence instead of the UniProt isoform and generated predicted structures with AlphaFold3.

The resulting table maps 72,745,436 possible missense variants in the human genome to amino acid substitutions in 19,576 proteins (UniProt ID), of which 18,968 are canonical and reviewed. This covers 19,678 of the 19,777 Ensembl gene IDs in the initial annotation table generated by gnomAD.

For proteins for which the UniProt isoform was available in the AlphaFold Protein Structure Database (AF2DB), we downloaded the corresponding PDB and PAE files. For proteins divided into multiple windows on AF2DB (longer than 2700 amino acids), PAE files were unavailable (190 UniProt IDs). For the sequences for which a structure was not available in AF2DB, we ran the AlphaFold 3 web server (AF3) to generate PDB and PAE files. We excluded a protein with only two amino acids (P0DPR3). AF3 has a limit of 5000 amino acids, so protein sequences that exceeded this were split into multiple windows. Each window was 5000 amino acids long, and windows overlapped by 1200 amino acids. For example, a protein of length 9000 would be divided into 3 windows: residues 1-5000, 3801-8800, and 7601-9000. The distance computation, described below, takes as input either a single PDB file or a set of PDB files for overlapping windows. AF3 generates 5 models, and we selected the model with the highest ranking score. . We used a total of 18,866 proteins from AF2DB and generated structures for 710 proteins in AF3. This resulted in a total of 21,219 PDB files from AF2DB and 728 from AF3.

We created an HDF5 table mapping all missense variants to UniProt ID, reference amino acid, alternate amino acid, position, and pdb file, for fast lookup. The HDF5 table, PDB files, PAE files, and metadata files are publicly available at https://tinyurl.com/22ue4ckk.

### Computing case and control allele counts in all neighborhoods

The 3DNT takes as input a set of genetic variants with allele count in cases and allele count in controls. By default, the software removes any variant with total allele count greater than five, that has allele frequency greater than 1e-3 in any population in gnomAD, or that falls into a low complexity region as annotated in UniProt. We map the variants to protein coordinates using the HDF5 table described above and create two L x 1-dimensional vectors, where L is the length of the protein: one for allele counts in cases and one for allele counts in controls. We drop any protein that does not have at least five case and five control mutations. We create n_sim_ = 1,000 null permutations of the data by drawing from the corresponding hypergeometric distribution (equivalent to permuting the case and control labels) and append these to the allele count vectors, resulting in case and control matrices of allele counts with dimension L x (1 + n_sim_).

To compute allele counts in neighborhoods, we first use the PDB files to construct distance matrices, representing each residue with its C_alpha_ atom. For large proteins with separate PDB files for multiple overlapping windows, the *i,j*-th entry of the distance matrix—which is used to determine whether *j* will be in the neighborhood centered at *i*—is computed in the PDB file in which *i* is closest to the center of the sequence, with distances set to infinity if there is no window containing both *i* and *j*. We binarize this matrix by thresholding at 15 angstroms. For proteins for which PAE (predicted aligned error) information is available (all proteins except those downloaded from the AlphaFold Database that required multiple overlapping windows), we additionally require the PAE to be less than 15. We then compute allele counts in neighborhoods by right multiplying by the per-residue allele count matrices.

### Significance testing

For each protein, we pre-compute a matrix of p-values for all possible pairwise case and control allele counts, up to the maximum case and control counts in the neighborhood count matrices. To do this efficiently, we first compute chi-square p-values for all pairs. Because we are only interested in small p-values, we set the final p-value equal to one wherever the chi-square p-value is greater than 0.1. For the remaining entries, we compute precise p-values with Fisher’s exact test. We use this matrix to look up p-values for every neighborhood in the protein, for the real data and the simulated data, by indexing into it with the neighborhood allele count matrices.

We conduct a large number of highly correlated tests. To correct for this, we use our null simulations to compute an empirical family-wise error rate (FWER) and false discovery rate (FDR). Our FWER for a raw p-value of P is computed in the standard way, as a ratio where the numerator is the number of null simulations with at least one neighborhood more significant than P and the denominator is the total number of null simulations. We compute empirical FDRs using the approach described in ref [52]. The false discovery rate at a given p-value threshold is equal to the expected number of false discoveries at that threshold divided by the total number of discoveries at that threshold. We estimate the expected number of false discoveries at threshold P_T_ by taking the average across the 1,000 null simulations of the number of neighborhoods with P≤P_T_ in the simulation. The total number of discoveries at P_T_ is the number of neighborhoods with P≤P_T_ in the real data. The FDR at P_T_ is

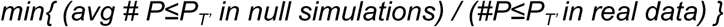

where the minimum is taken over P_T’_>P_T_, reducing noise and ensuring that FDR is monotonic in p-value.

### Choice of radius

In choosing the radius for the 3DNT, there are two competing considerations. A smaller radius increases interpretability, while a larger radius allows for more variants in the neighborhood, increasing power at a given effect size. To balance these, we used the SCHEMA data to estimate the average number of ultra-rare case and control variants per neighborhood at a given radius, and the odds ratio in that neighborhood that would be required to achieve a p-value of 1e-5 (Table S1). We found that 15 angstroms was the smallest radius at which an odds ratio below five would result in P<1e-5 in an average-sized neighborhood in an average-sized protein. A radius of 10, on the other hand, would require an odds ratio of 16, which we would be unlikely to see.

### PAE filtering

To assess whether filtering on PAE improved the accuracy of the method, we ran our analysis of ClinVar data both with and without a PAE filter and measured the average log distance of the top neighborhood centers to the nearest UniProt-annotated active or binding site. Top neighborhoods are defined as the most significant neighborhood per protein with FDR < 0.05. We found that top neighborhood distances to active and binding sites were shorter on average when using the PAE filter, though not significantly so (P=0.9). We thus incorporated this filter into the method by default, but we do not expect the filter to have a large impact on the results.

### ClinVar data

ClinVar variant data were downloaded from the NCBI FTP server (hg38 VCF, clinvar.vcf.gz). Variants were restricted to SNVs on autosomes and chromosome X. Variants were assigned a case allele count of one and a control allele count of zero if the aggregate clinical significance (CLNSIG) was Pathogenic, Likely pathogenic, or Pathogenic/Likely pathogenic. Variants were assigned a control allele count of one and a case allele count of zero if CLNSIG was Benign, Likely benign, or Benign/Likely benign. All other variants were excluded. At least one ClinVar review star was required. Additional variants with a control allele count of one and a case allele count of zero were added if they were common in at least one population according to gnomAD, with common being defined as an allele count of at least 100 and an allele frequency greater than 1e-3. Lists of autosomal dominant and autosomal recessive disease genes were derived from the Human Phenotype Ontology (HPO) gene-phenotype annotation file (genes_to_phenotype.txt), using HPO terms HP:0000006 (autosomal dominant inheritance) and HP:0000007 (autosomal recessive inheritance), respectively.

### Data from the Autism Sequencing Consortium

The Autism Sequencing Consortium (ASC) aggregated trio data from 37,486 probands with autism and 9,567 unaffected siblings, with all variants capped at a 0.1% maximum population frequency. Data collection and processing is described in ref [29]. We assigned the “case variant” label to *de novo* variants from the probands and the “control variant” label to singleton untransmitted variants (that is, parental alleles with a transmitted count of 0 and an untransmitted count of 1). We do not include *de novo* variants from an additional 1,194 probands analyzed by the ASC for whom inherited data was not available.

### Case-control exome sequencing data sets

We downloaded public data from the SCHEMA browser (https://schema.broadinstitute.org) and Epi25 browser (https://epi25.broadinstitute.org/). In both cases, we used Hail (https://hail.is) to liftover from GRCh37 to GRCh38. We removed variants not measured in all cases and all controls.

### Characterization of top neighborhoods in ClinVar

To assess whether significant neighborhoods were enriched near known structural features, we compared the locations of the most significant neighborhoods with respect to structural features against the locations of randomly selected neighborhoods. For each protein with at least one neighborhood reaching FDR < 0.05, we selected the most significant neighborhood; for comparison, we selected a random neighborhood from each remaining protein.

We evaluated three structural features: (1) burial, quantified by relative solvent accessible surface area (RSASA) computed using the get_sasa_relative function in PyMOL, with residues below RSASA = 0.25 classified as buried; (2) UniProt-annotated active and binding sites, as formatted in the G2P portal^53^; and (3) predicted small molecule binding pockets from AF2Bind^33^. For burial, we computed a chi-square test to determine if the centers of significant neighborhoods are more likely to be buried than the centers of random neighborhoods. For the remaining features, we computed the shortest distance from each neighborhood center to the nearest annotated residue, then compared distances between the significant and random sets using a t-test on log-transformed distances. Proteins lacking active or binding site annotations were assigned a default distance of 100 Å; AF2Bind comparisons were restricted to proteins present in that dataset.

For evaluating the proportion of case variants in the hydrophobic core, we first computed, for each protein with a significant neighborhood and each randomly selected protein, the proportion of P/LP variants that were in the hydrophobic core, and then computed a two-sided t-test between the proportions in significant neighborhoods and random neighborhoods. We defined hydrophobic core variants as variants with RSASA<0.1, a hydrophobic reference amino acid (one of A, I, L, M, V, F, W, Y), and a hydrophilic alternate amino acid (not in the previous list). Selected neighborhoods were restricted to those with at least one case variant for this analysis.

### AlphaMissense-based neighborhoods

For every gene, we defined synthetic “case” mutations as those with AM > 0.99 (or the top 500 if more than 500 exceeded this threshold) and synthetic “control” mutations as a sample from the bottom 50% of AM scores (up to 1,000 mutations). With these definitions, we ran 3DNT genome-wide.

### FoldX analysis of KCNA1 G374R

We first generated an AlphaFold3-predicted structure of the KCNA1/KCNA2 heterotetramer in complex with K⁺. Using FoldX, we calculated the change in binding energy (ΔΔG) associated with the mutation, following the protocol described in ref [54]. Briefly, the protein complex structure was refined by running the “RepairPDB” function five times. The G374R mutation was then introduced using the “BuildModel” function, generating 10 independent mutant models. For each model, the change in interaction energy was computed with “AnalyseComplex”, and ΔΔG values were averaged across the 10 runs. We calculated average changes in interaction energy for multiple chain-chain combinations within the complex. We observed no significant change in binding affinity between KCNA1 and K⁺. In contrast, the ΔΔG between KCNA1 and KCNA2 subunits exceeded 25 kcal/mol, indicating a potentially substantial destabilization of their interaction. These results suggest that the G374R mutation may not primarily impact potassium binding. Rather, it likely disrupts interactions between the KCNA1 and KCNA2 subunits, destabilizing heterotetramer formation itself.

### PSCAN Neighborhoods

PSCAN and 3DNT define neighborhoods quite differently. For each distance threshold, PSCAN creates a graph with nodes representing residues and edges connecting residue pairs within that distance threshold. For each graph, it defines a set of neighborhoods corresponding to the set of connected components in the graph. It then takes all neighborhoods from the graphs defined at all distance thresholds (a finite set of graphs despite the infinite set of potential distance thresholds). The resulting set of neighborhoods is very different from the set of spherical neighborhoods tested in 3DNT. To understand whether PSCAN’s failure to identify enrichments near functional sites (**Figure 5**) might be due to its neighborhood definition procedure, we took the list of UniProt-annotated active and binding sites, proteome-wide. We chose 1,000 sites at random, and for each of these sites, we computed the minimum jaccard distance of the sphere centered at the site to any PSCAN neighborhood in the same protein, using fixed radii of 10 and 15 angstroms. For both radii, we found that only 8% of sites had a PSCAN neighborhood with Jaccard distance < 0.5 (Figure S1). These results suggest that PSCAN neighborhoods are not well-suited to capturing the compact signals at functional sites that 3DNT is designed to detect.

To repurpose PSCAN into a method that identifies 3D neighborhoods that are enriched relative to a gene-specific background, we gave it case and control allele counts within a gene rather than the total case and control counts for the study as the input to use for the null distribution. To apply it to our data of case and control counts, we converted the counts into the score statistics required as input to PSCAN, using the same assumptions (no LD and no relevant covariates) that are used in 3DNT.

### Analysis with POINT

To compare POINT to 3DNT, we made four adaptations. First, as with PSCAN, we analyzed one gene at a time, to identify neighborhoods with enrichment relative to the gene and to avoid finding genes with disease association but without spatial structure. Second, POINT does not include any automated mapping of variants to 3D structures, so we used infrastructure from 3DNT for this. Third, POINT requires individual level data, so we took the assumptions of 3DNT (no linkage disequilibrium, no relevant covariates) and constructed pseudo-individual data from allele counts. Fourth, because we apply POINT separately to each gene to make the test competitive within-gene, we corrected for the total number of tests by combining all p-values and using standard Benjamini Hochberg and Bonferroni corrections.

With these adaptations, POINT successfully ran on our data. However, there were no significant results. In the manuscript describing POINT, the successful applications included larger allele counts, while our application involved single-gene analyses with all minor allele counts under five. We hypothesize that the statistical approximations made may not be relevant in our data regime; however, further work will be needed.

### The Autism Sequencing Consortium (ASC)

Branko Aleksic^42^, Mykyta Artomov^7,8,11,43^, Chiara Auwerx^7,8,10,11^, Mafalda Barbosa^3,6^, Elisa Benetti^44,45^, Catalina Betancur^41^, Monica Biscaldi-Schafer^46^, Anders D. Børglum^47,48,49,50^, Harrison Brand^7,11,43,51^, Alfredo Brusco^52,53^, Joseph D. Buxbaum^1,2,3,4,5,6^, Gabriele S. Campos^14^, Simona Cardaropoli^54^, Diana Carli^54^, Angel Carracedo^55,56^, Marcus C. Y. Chan^57^, Andreas G. Chiocchetti^42^, Brian H. Y. Chung^57^, Brett Collins^1,2,6^, Ryan L. Collins^7,11,33,43^, Edwin H. Cook^38^, Hilary Coon^58,59^, Claudia I. S. Costa^14^, Michael L. Cuccaro^17,18^, David J. Cutler^60^, Mark J. Daly^7,8,9,10,34,35^, Silvia De Rubeis^1,2,4,6,29^, Bernie Devlin^12^, Ryan N. Doan^61^, Enrico Domenici^62^, Shan Dong^63^, Chiara Fallerini^44,45^, Magdalena Fernandez^16^, Montserrat Fernández-Prieto^55,64^, Giovanni Battista Ferrero^54^, Eugenio Ferro^16^, Jennifer Foss-Feig^1,2,6^, Christine M. Freitag^42^, Jack M. Fu^7,10,11^, Liliana Galeano^19^, J. Jay Gargus^65^, Sherif Gerges^7,8,11,43^, Elisa Giorgio^52^, Ana Cristina D. E. S. Girardi^14^, Stephen Guter^66^, Emily Hansen-Kiss^67^, Erina Hara^1,2^, Gail E. Herman^68^, Luis C. Hernandez^19^, Irva Hertz-Picciotto^20^, David M. Hougaard^46,69^, Christina M. Hultman^70^, Suma Jacob^66^, Miia Kaartinen^71^, Lambertus Klei^12^, Alexander Kolevzon^1,2,30^, Itaru Kushima^47,72^, Maria C. Lattig^19^, So Lun Lee^57^, Terho Lehtimäki^73^, Lindsay Liang^63^, Carla Lintas^74^, Alicia Ljungdahl^63^, Andrea del Pilar Lopez^15^, Caterina Lo Rizzo^44,45^, Yunin Ludena^20^, Patricia Maciel^75^, Behrang Mahjani^1,2,3,6,36,37^, Nell Maltman^66^, Marianna Manara^45,76^, Dara S. Manoach^77^, Gal Meiri^78,79^, Idan Menashe^80,81^, Judith Miller^82,83^, Nancy Minshew^12^, Matthew Mosconi^84^, Marina Natividad Avila^1,2,3,4,5,6^, Rachel Nguyen^65^, Norio Ozaki^47,85^, Aarno Palotie^7,9,35,86^, Mara Parellada^87^, Maria Rita Passos-Bueno^14^, Lisa Pavinato^52^, Katherine P. Peña^19^, Minshi Peng^88^, Margaret Pericak-Vance^17,18^, Antonio M. Persico^89^, Isaac N. Pessah^20^, Thariana Pichardo^1,2,4,6^, Kaija Puura^71^, Abraham Reichenberg^1,2,6,90^, Alessandra Renieri^44,45,76^, Kathryn Roeder^39,40^, Catherine Sancimino^1,2^, Stephan J. Sanders^91,92,93^, Sven Sandin^1,2,70^, F. Kyle Satterstrom^7,8,9^, Stephen W. Scherer^94,95^, Sabine Schlitt^42^, Rebecca J. Schmidt^20^, Lauren Schmitt^66^, Katja Schneider-Momm^42^, Paige M. Siper^1,2,6^, Laura Sloofman^1,2,3,4,5,6^, Moyra Smith^65^, Renee Soufer^1,2^, Christine R. Stevens^7,8,9^, Pål Suren^96^, James S. Sutcliffe^97,98^, John A. Sweeney^99^, Michael E. Talkowski^7,8,10,11,33^, Flora Tassone^20,31^, Karoline Teufel^42^, Elisabetta Trabetti^100^, Slavica Trajkova^52^, M. Pilar Trelles^32^, Brie Wamsley^101^, Jaqueline Y. T. Wang^14^, Lauren A. Weiss^63^, Mullin H. C. Yu^57^, Ryan Yuen^94^

^1^Seaver Autism Center for Research and Treatment, Icahn School of Medicine at Mount Sinai, New York, New York, USA. ^2^Department of Psychiatry, Icahn School of Medicine at Mount Sinai, New York, New York, USA. ^3^Department of Genetics and Genomic Sciences, Icahn School of Medicine at Mount Sinai, New York, New York, USA. ^4^Friedman Brain Institute, Icahn School of Medicine at Mount Sinai, New York, New York, USA. ^5^Department of Neuroscience, Icahn School of Medicine at Mount Sinai, New York, New York, USA. ^6^The Mindich Child Health and Development Institute, Icahn School of Medicine at Mount Sinai, New York, New York, USA. ^7^Program in Medical and Population Genetics, Broad Institute of MIT and Harvard, Cambridge, Massachusetts, USA. ^8^Stanley Center for Psychiatric Research, Broad Institute of MIT and Harvard, Cambridge, Massachusetts, USA. ^9^Analytic and Translational Genetics Unit, Department of Medicine, Massachusetts General Hospital, Boston, Massachusetts, USA. ^10^Center for Genomic Medicine, Department of Medicine, Massachusetts General Hospital, Boston, Massachusetts, USA. ^11^Department of Neurology, Massachusetts General Hospital and Harvard Medical School, Boston, Massachusetts, USA. ^12^Department of Psychiatry, University of Pittsburgh School of Medicine, Pittsburgh, Pennsylvania, USA. ^13^Division of Research, Kaiser Permanente Northern, Pleasanton, California, USA. ^14^Centro de Estudos do Genoma Humano e Células-Tronco, Departamento de Genética e Biologia Evolutiva, Instituto de Biociências, Universidade de São Paulo, São Paulo, Brasil. ^15^Facultad de Medicina, Universidad de los Andes, Bogotá, Colombia. ^16^Instituto Colombiano del Sistema Nervioso, Clínica Montserrat, Bogotá, Colombia. ^17^John P. Hussman Institute for Human Genomics, University of Miami Miller School of Medicine, Miami, Florida, USA. ^18^The Dr. John T. Macdonald Foundation Department of Human Genetics, University of Miami Miller School of Medicine, Miami, Florida, USA. ^19^Facultad de Ciencias, Universidad de los Andes, Bogotá, Colombia. ^20^MIND (Medical Investigation of Neurodevelopmental Disorders) Institute, University of California Davis, Davis, California, USA. ^21^Department of Psychiatry, Yale University School of Medicine, New Haven, Connecticut, USA. ^22^National Center of Posttraumatic Stress Disorders, VA CT Healthcare Center, West Haven, Connecticut, USA. ^23^Centro Ann Sullivan del Peru, Lima, Peru. ^24^Center Ann Sullivan International, Lawrence, Kansas, USA. ^25^Hospital Psiquiátrico Infantil Dr. Juan N. Navarro, Ciudad de México, Mexico. ^26^Universidad Nacional Autónoma de México, Ciudad de México, Mexico. ^27^Kaiser Permanente School of Medicine, Pasadena, California, USA. ^28^Departamento de Genética, Subdirección de Investigaciones Clínicas, Instituto Nacional de Psiquiatría Ramón de la Fuente Muñiz México, Ciudad de México, Mexico. ^29^The Alper Center for Neural Development and Regeneration, Icahn School of Medicine at Mount Sinai, New York, New York, USA. ^30^Department of Pediatrics, Icahn School of Medicine at Mount Sinai, New York, New York, USA. ^31^Department of Biochemistry and Molecular Medicine, University of California Davis, School of Medicine, Davis, California, USA. ^32^Psychiatry and Behavioral Sciences, Boston Children’s Hospital, Boston, Massachusetts, USA. ^33^Program in Bioinformatics and Integrative Genomics, Harvard Medical School, Boston, Massachusetts, USA. ^34^Department of Medicine, Harvard Medical School, Boston, Massachusetts, USA. ^35^Institute for Molecular Medicine Finland (FIMM), University of Helsinki, Helsinki, Finland. ^36^Department of Artificial Intelligence and Human Health, Icahn School of Medicine at Mount Sinai, New York, New York, USA. ^37^Department of Molecular Medicine and Surgery, Karolinska Institutet, Stockholm. ^38^Department of Psychiatry, University of Illinois Chicago, Chicago, Illinois,

USA. ^39^Department of Statistics, Carnegie Mellon University, Pittsburgh, Pennsylvania, USA. ^40^Computational Biology Department, Carnegie Mellon University, Pittsburgh, Pennsylvania, USA. ^41^Sorbonne Université, INSERM, CNRS, Institut de Biologie Paris Seine, Center for Neuroscience at Sorbonne Université, Paris, France. ^42^Department of Psychiatry, Graduate School of Medicine, Nagoya University, Nagoya, Japan. ^43^Center for Genomic Medicine, Massachusetts General Hospital, Boston, Massachusetts, USA. ^44^Med Biotech Hub and Competence Center, Department of Medical Biotechnologies, University of Siena, Siena, Italy. ^45^Medical Genetics, University of Siena, Siena, Italy. ^46^Department of Child and Adolescent Psychiatry, Psychosomatics and Psychotherapy, Goethe University Frankfurt, Frankfurt, Germany. ^47^The Lundbeck Foundation Initiative for Integrative Psychiatric Research, iPSYCH, Aarhus, Denmark. ^48^Department of Biomedicine—Human Genetics, Aarhus University, Aarhus, Denmark. ^49^Center for Genomics and Personalized Medicine, Aarhus, Denmark. ^50^Bioinformatics Research Centre, Aarhus University, Aarhus, Denmark. ^51^Pediatric Surgical Research Laboratories, Department of Surgery, Massachusetts General Hospital, Boston, Massachusetts, USA. ^52^Department of Medical Sciences, University of Torino, Turin, Italy. ^53^Medical Genetics Unit, ‘Città della Salute e della Scienza’ University Hospital, Turin, Italy. ^54^Department of Public Health and Pediatrics, University of Torino, Turin, Italy. ^55^Grupo de Medicina Xenómica, Centro de Investigación en Red de Enfermedades Raras (CIBERER), CIMUS, Universidade de Santiago de Compostela, Santiago de Compostela, Spain. ^56^Fundación Pública Galega de Medicina Xenómica, Servicio Galego de Saúde (SERGAS), Santiago de Compostela, Spain. ^57^Department of Pediatrics and Adolescent Medicine, Duchess of Kent Children’s Hospital, The University of Hong Kong, Hong Kong Special Administrative Region, China. ^58^Department of Internal Medicine, University of Utah, Salt Lake City, Utah, USA. ^59^Department of Psychiatry, Huntsman Mental Health Institute, University of Utah, Salt Lake City, Utah, USA. ^60^Department of Human Genetics, Emory University School of Medicine, Atlanta, Georgia, USA. ^61^Division of Genetics and Genomics, Boston Children’s Hospital, Boston, Massachusetts, USA. ^62^Department of Cellular, Computational and Integrative Biology, University of Trento, Trento, Italy. ^63^Department of Psychiatry, UCSF Weill Institute for Neurosciences, University of California San Francisco, San Francisco, California, USA. ^64^Neurogenetics group, Instituto de Investigación Sanitaria de Santiago (IDIS-SERGAS), Santiago de Compostela, Spain. ^65^Center for Autism Research and Translation, University of California Irvine, Irvine, California, USA. ^66^Institute for Juvenile Research, Department of Psychiatry, University of Illinois at Chicago, Chicago, Illinois, USA. ^67^Department of Diagnostic and Biomedical Sciences, University of Texas Health Science Center at Houston, School of Dentistry, Houston, Texas, USA. ^68^The Research Institute at Nationwide Children’s Hospital, Columbus, Ohio, USA. ^69^Center for Neonatal Screening, Department for Congenital Disorders, Statens Serum Institut, Copenhagen, Denmark. ^70^Department of Medical Epidemiology and Biostatistics, Karolinska Institutet, Stockholm, Sweden. ^71^Department of Child Psychiatry, Tampere University and Tampere University Hospital, Tampere, Finland. ^72^Medical Genomics Center, Nagoya University Hospital, Nagoya, Japan. ^73^Department of Clinical Chemistry, Fimlab Laboratories and Finnish Cardiovascular Research Center-Tampere, Faculty of Medicine and Health Technology, Tampere University, Tampere, Finland. ^74^Service for Neurodevelopmental Disorders, University Campus Bio-medico of Rome, Rome, Italy. ^75^Life and Health Sciences Research Institute, School of Medicine, University of Minho, Braga, Portugal. ^76^Genetica Medica, Azienda Ospedaliera Universitaria Senese, Siena, Italy. ^77^Department of Psychiatry, Massachusetts General Hospital and Harvard Medical School, Boston, Massachusetts, USA. ^78^The Azrieli National Center for Autism and Neurodevelopment Research, Ben-Gurion University of the Negev, Beer-Sheva, Israel. ^79^Pre-School Psychiatry Unit, Soroka University Medical Center, Beer Sheva, Israel. ^80^Department of Public Health, Ben-Gurion University of the Negev, Beer-Sheva, Israel. ^81^National Autism Research Center of Israel, Ben-Gurion University of the Negev, Beer-Sheva, Israel. ^82^Children’s Center for Autism Research and Training, University of Kansas, Lawrence, Kansas, USA. ^83^Department of Psychiatry, University of Utah, Salt Lake City, Utah, USA. ^84^Life Span Institute and Kansas Center for Autism Research and Training, University of Kansas, Lawrence, Kansas, USA. ^85^Institute for Glyco-core Research (iGCORE), Nagoya University, Nagoya, Japan. ^86^Psychiatric & Neurodevelopmental Genetics Unit, Department of Psychiatry, Massachusetts General Hospital, Boston, Massachusetts, USA. ^87^Department of Child and Adolescent Psychiatry, Hospital General Universitario Gregorio Marañón, IiSGM, CIBERSAM, School of Medicine Complutense University, Madrid, Spain. ^88^Department of Statistics and Data Science, Carnegie Mellon University, Pittsburgh, Pennsylvania, USA. ^89^Interdepartmental Program ‘Autism 0-90’, ‘Gaetano Martino’ University Hospital, University of Messina, Messina, Italy. ^90^Department of Environmental Medicine and Public Health, Icahn School of Medicine at Mount Sinai, New York, New York, USA. ^91^Institute of Developmental and Regenerative Medicine, Department of Paediatrics, University of Oxford, Oxford, UK ^92^Department of Psychiatry and Behavioral Sciences, UCSF Weill Institute for Neurosciences, University of California, San Francisco, San Francisco, California, USA. ^93^New York Genome Center, New York, New York, USA. ^94^Program in Genetics and Genome Biology, The Centre for Applied Genomics, The Hospital for Sick Children, Toronto, Ontario, Canada. ^95^Department of Molecular Genetics and McLaughlin Centre, University of Toronto, Toronto, Ontario, Canada. ^96^Norwegian Institute of Public Health, Oslo, Norway. ^97^Department of Molecular Physiology & Biophysics and Psychiatry, Vanderbilt University School of Medicine, Nashville, Tennessee, USA. ^98^Vanderbilt Genetics Institute, Vanderbilt University School of Medicine, Nashville, Tennessee, USA. ^99^Department of Psychiatry, University of Cincinnati, Cincinnati, Ohio, USA. ^100^Department of Neurosciences, Biomedicine and Movement Sciences, Section of Biology and Genetics, University of Verona, Verona, Italy. ^101^Program in Neurogenetics, Department of Neurology, David Geffen School of Medicine, University of California Los Angeles, Los Angeles, California, USA.

---

**Figure S1:**
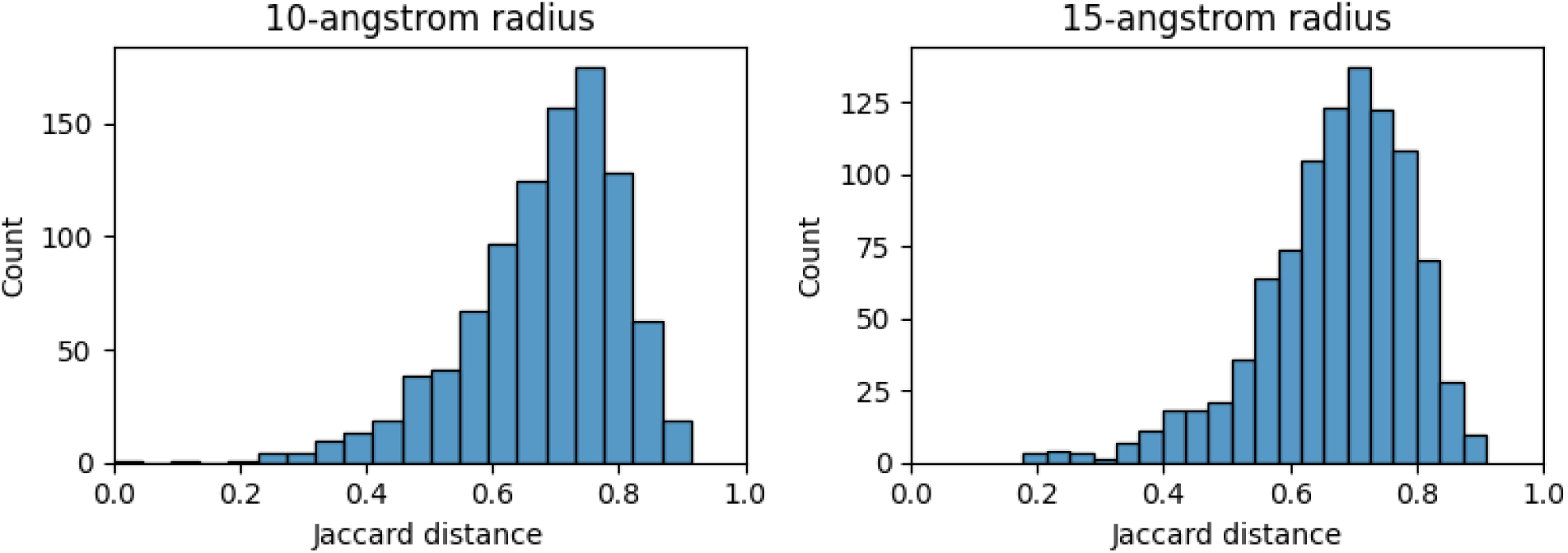
For 1,000 uniformly randomly chosen active or binding sites, the minimum Jaccard distance to any PSCAN neighborhood of a sphere centered at the site.

**Table S1.** Odds ratio required in an average-sized neighborhood in an average-sized protein to achieve P<1e-5.

| Radius (angstroms) | Odds ratio |
| --- | --- |
| 10 | 15.77 |
| 11 | 9.31 |
| 12 | 8.07 |
| 13 | 5.73 |
| 14 | 5.16 |
| 15 | 4.59 |
| 16 | 4.37 |
| 17 | 4.07 |
| 18 | 3.62 |
| 19 | 3.48 |
| 20 | 3.36 |

